# Patterns of compliance with COVID-19 preventive behaviours: a latent class analysis of 20,000 UK adults

**DOI:** 10.1101/2021.03.16.21253717

**Authors:** Liam Wright, Andrew Steptoe, Daisy Fancourt

## Abstract

**Background:** Governments have implemented a range of measure to tackle COVID-19, primarily focusing on changing citizens’ behaviours in order to lower transmission of the virus. Few studies have looked at the patterns of compliance with different measures within individuals: whether people comply with all measures or selectively choose some but not others. Such research is important for designing interventions to increase compliance.

**Methods:** We used cross-sectional data from 20,947 UK adults in the COVID-19 Social Study collected 17 November – 23 December 2020. Self-report compliance was assessed with six behaviours: mask wearing, hand washing, indoor household mixing, outdoor household mixing, social distancing, and compliance with other guidelines. Patterns of compliance behaviour were identified using latent class analysis, and multinomial logistic regression was used to assess demographic, socioeconomic and personality predictors of behaviour patterns.

**Results:** We selected a four latent class solution. Most individuals reported similar levels of compliance across the six behaviour measures. High levels of compliance was the modal response. Lower self-reported compliance was related to young age, high risk-taking behaviour, low confidence in government, and low empathy, among other factors. Looking at individual behaviours, mask wearing had the highest level of compliance whilst compliance with social distancing was relatively low.

**Conclusion:** Results suggest that individuals choose to comply with all guidelines, rather than some but not others. Strategies to increase compliance should focus on increasing general motivations to comply alongside specifically encouraging social distancing.

## Introduction

Governments have implemented a series of measures to tackle the spread of COVID-19. Many of these measures have focused on changing citizens’ behaviours, such as advertising personal hygiene reminders (e.g., washing hands), mandating the wearing of face masks, recommending social distancing in public spaces, and prohibiting household mixing. These interventions are effective at reducing the spread of the virus (Chu et al., 2020) but require voluntary cooperation on behalf of citizens. Compliance with these behaviours is not complete (Ipsos MORI, 2021; YouGov, 2020).

Given the importance of these measures for tackling pandemics, a sizeable literature has emerged on the determinants and predictors of compliance, both during the current pandemic (Perra, 2021) and from previous epidemics (Bish & Michie, 2010). Many of these studies have focused on specific behaviours (Bargain & Aminjonov, 2020) or on compliance with guidelines in general (Wright et al., Accepted; Wright & Fancourt, 2020). However, an understudied area is how compliance behaviours “cluster” within individuals; for instance, whether individuals perform some behaviours and not others. Understanding patterns of compliance is important as it may reveal information on behaviours individuals find particularly difficult.

Studies from the current pandemic (Brouard et al., 2020; Kamenidou et al., 2020; Tomczyk et al., 2020; Toussaint et al., 2020) and previous epidemics (Liao et al., 2015) using a range of methodological approaches, including cluster analysis (Kamenidou et al., 2020), factor analysis (Breakwell et al., 2021; Brouard et al., 2020; Toussaint et al., 2020), and latent class analysis (Liao et al., 2015; Tomczyk et al., 2020), have found several distinct patterns in compliance behaviours (though, also see, Breakwell et al., 2021). For instance, a recent German study identified a small group of individuals intending to comply only with behaviours performed in public, such as social distancing and avoiding mass events (Tomczyk et al., 2020). However, existing studies are typically based on small samples, use data from early in the COVID-19 pandemic (which is limiting as predictors differ through time; van der Weerd et al., 2011; Wright & Fancourt, 2020), or have looked at compliance intentions, rather than performed behaviours.

Therefore, in this study we explored patterns on self-reported compliance with six COVID-19 preventive behaviours, using data from a sample of 20,000 UK adults eight months after lockdown was first implemented in the UK. We further tested whether behavioural patterns were related to a wide-range of demographic, socio-economic and personality trait characteristics.

## Methods

### Participants

We used data from the COVID-19 Social Study; a large panel study of the psychological and social experiences of over 70,000 adults (aged 18+) in the UK during the COVID-19 pandemic. The study commenced on 21 March 2020 and involved online weekly (from August 2020, monthly) data collection across the pandemic in the UK. The study is not random and therefore is not representative of the UK population, but it does contain a heterogeneous sample. The sample was recruited using three primary approaches. First, convenience sampling was used, including promoting the study through existing networks and mailing lists (including large databases of adults who had previously consented to be involved in health research across the UK), print and digital media coverage, and social media. Second, more targeted recruitment was undertaken focusing on (i) individuals from a low-income background, (ii) individuals with no or few educational qualifications, and (iii) individuals who were unemployed. Third, the study was promoted via partnerships with third sector organisations to vulnerable groups, including adults with pre-existing mental health conditions, older adults, carers, and people experiencing domestic violence or abuse. The study was approved by the UCL Research Ethics Committee [12467/005] and all participants gave informed consent. The study protocol and user guide (which includes full details on recruitment, retention, data cleaning, weighting and sample demographics) are available at https://github.com/UCL-BSH/CSSUserGuide.

A module on compliance behaviours was included in the survey between 17 November and 23 December 2020. For these analyses, we focused on individuals with complete observed compliance behaviours (n = 21,066; 91.6% of individuals interviewed between these dates). We excluded participants with missing data on key demographic data that we used to construct survey weights (n = 119), leaving a sample of 20,947. This sample represents 29.5% of those with data collection by 23 December 2020.

The period 17 November to 23 December 2020 overlaps with the second wave of COVID-19 in the UK in which there were several changes to COVID-19 related rules. Description of changes to these rules is provided in the Supplementary Information. Supplementary Figure S1 shows 7-day COVID-19 caseloads and confirmed deaths, along with the Oxford Policy Tracker, a numerical summary of policy stringency (Hale et al., 2020), across the study period.

### Measures

#### Compliance Behaviours

We analysed six compliance behaviours: hand washing, face mask wearing, social distancing, household mixing indoors, household mixing outdoors, and compliance with other guidelines. Participants were asked for their compliance with these behaviours over the previous seven days (see Supplementary Information for question precise wording). The response categories were: never, rarely, occasionally, frequently, and always. Items on household mixing were phrased such that higher scores indicated lower compliance. We reverse coded these for consistency with the other behaviours.

#### Predictors of Compliance Behaviour

We assessed a range of demographic, socioeconomic, and personality trait characteristics as predictors of compliance behaviour. We selected these predictors using previous results from the COVID-19 Social Study (Wright & Fancourt, 2020) and considering the COM-B framework of health behaviour (Michie et al., 2011). The COM-B model posits that behaviour is determined by (subjective and objective) capability, opportunity for action, and autonomic and reflective motivation.

For capability to comply, we included variables for locus of control, neighbourhood crowding, annual household income, educational level, diagnosed psychiatric condition, and ethnicity. For opportunity to comply, we included variables for lockdown tier, country of residence, household income, date of data collection (natural splines, 2 degrees of freedom). For motivation to comply, we include variables for long-term physical health conditions (0, 1, 2+), age (gender), sex, keyworker status, self-isolation during first data collection, Big-5 personality traits (openness, conscientiousness, extraversion, agreeableness, and neuroticism), trait empathy, risk-taking, household overcrowding (1+ persons per room), living arrangement (alone, not alone without child, not alone with child), confidence in government, and mental health experiences during the first lockdown (same, better or worse vis-à-vis prior to the pandemic). Many of these variables were measured at baseline data collection or in modules contained in the survey prior to measurement of compliance. More detail on the measurement of the variables in this analysis is provided in the Supplementary Information.

### Statistical Analysis

We estimated latent class models using robust maximum likelihood estimation with 2,000 sets of random starting values, repeated for 1-9 latent classes. Compliance variables were treated as ordered categorical variables in these models. We selected the final model considering the Bayes Information Criterion (BIC) and entropy values, average latent class probabilities and substantive interpretation of the classes identified. We included probability weights in models and so were not able to use the bootstrapped likelihood ratio test for model selection.

After selecting a final latent class solution, we calculated multinomial logistic regression models to predict compliance patterns according to demographic, socioeconomic and psychological factors using the 3-step approach (Asparouhov & Muthén, 2014). Due to data missingness, we used multiple imputation in these models (40 imputed datasets, 10 iterations). Data were imputed using the mice R packages with continuous variables imputed using predictive mean matching and categorical variables imputed using multinomial logistic regression. Latent class probabilities were included as auxiliary variables in the imputation models.

We found that average compliance levels differed across the six compliance behaviours. Consequently, as a further analysis, we also estimated models of the predictors of individual compliance behaviours using ordinal regression. We again used multiple imputed data for these models, with separate imputations run for each compliance measure (40 datasets, 10 iterations). No auxiliary variables were included in these models.

In the regression results, we scaled continuous variables, such that a one-unit change is equivalent to 2 SD change in the variable to allow comparison with categorical variables. The code to replicate the analysis is available at https://osf.io/aswqc/.

## Results

### Descriptive Statistics

Sample descriptive statistics are shown in Supplementary Table S1. The distributions of the individual compliance behaviours are displayed in Figure 1. For each of the behaviours, the majority of participants reported frequent or complete compliance. Full compliance was greatest for mask wearing and indoor and outdoor social mixing. Lowest compliance levels were observed for hand washing and social distancing. Exploratory factor analysis of the six items identified two factors with eigenvalues greater than 1 (polychoric correlations; promax rotation). Items on mask wearing, other guidelines, hand washing, and social distancing loaded onto one factor and the two items on household mixing loaded onto the other (for factor loadings, see Supplementary Figure S2). The correlation between the factors was 0.41. Chronbach’s α was 0.61 for the six items.

**Figure 1:**
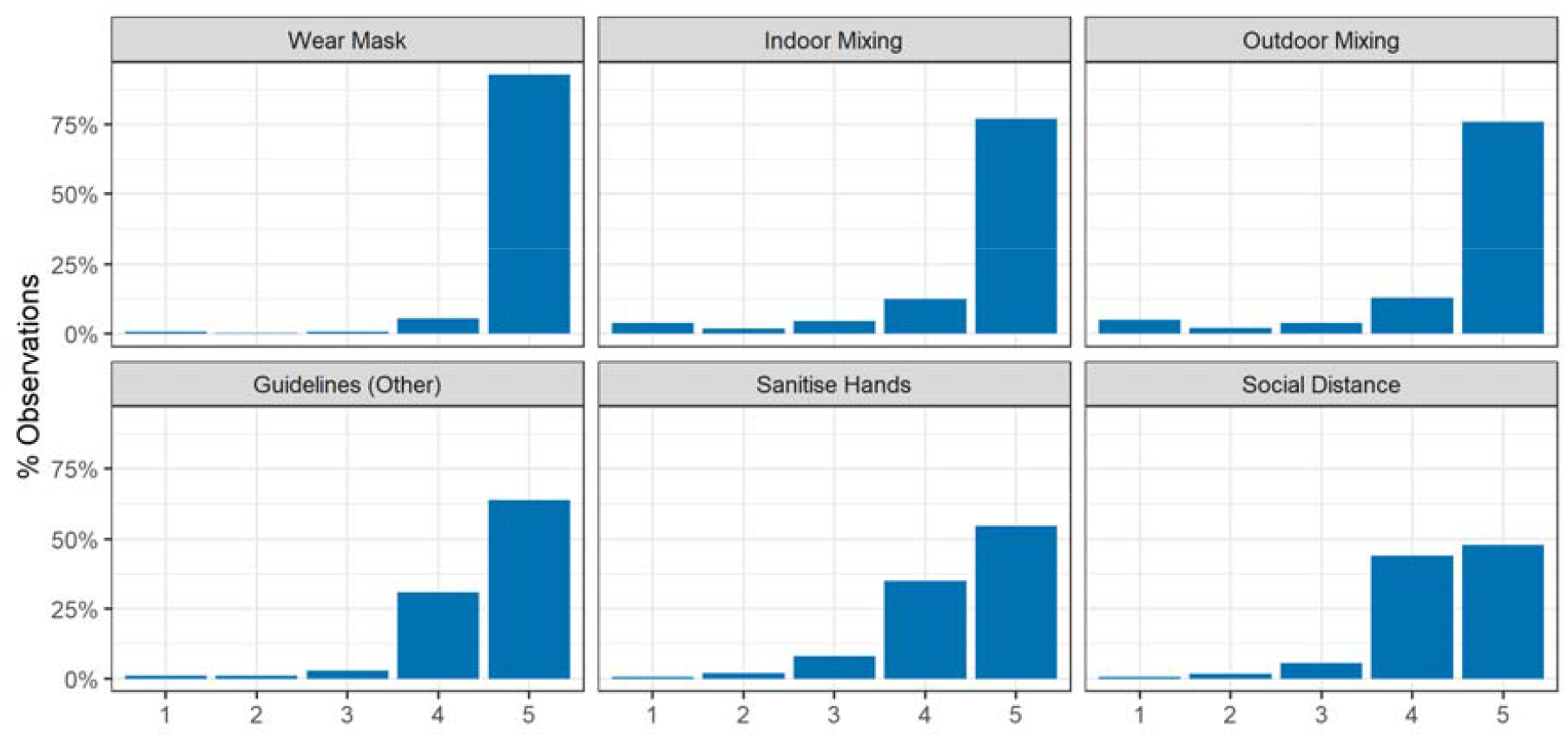
Distribution of compliance by behaviour. Response categories: 1. Never, 2, Rarely, 3. Occasionally, 4. Frequently, and 5. Always. Items on household mixing were phrased with higher values indicating lower compliance, and so are reverse coded for consistency with the other items.

### Patterns of Compliance Behaviour

Fit statistics for the latent class analysis models are displayed in Table 1. A BIC “elbow” plot is provided in Supplementary Figure S3. We selected a four-class solution as this had the highest entropy values (0.84) and the less parsimonious five-class solution provided substantively similar results. Mean values for each behaviour according to class for the four-class solution are displayed in Figure 2 (Supplementary Figure S4 alternatively displays this information as predicted probabilities). Sample sizes and last class probabilities are displayed in Table 2.

**Table 1:** Latent Class Analysis Fit Statistics

| Classes | AIC | BIC | Entropy | Average Class Probability |  |
| --- | --- | --- | --- | --- | --- |
|  |  |  |  | Min. | Max. |
| 1 | 201,155 | 201,346 |  | 1.000 | 1.00 |
| 2 | 184,808 | 185,198 | 0.75 | 0.900 | 0.94 |
| 3 | 179,920 | 180,508 | 0.84 | 0.870 | 0.97 |
| 4 | 176,180 | 176,967 | 0.82 | 0.811 | 0.95 |
| 5 | 174,645 | 175,631 | 0.77 | 0.770 | 0.95 |
| 6 | 173,956 | 175,141 | 0.79 | 0.767 | 0.97 |
| 7 | 173,317 | 174,700 | 0.80 | 0.678 | 0.97 |
| 8 | 172,981 | 174,563 | 0.81 | 0.678 | 0.97 |
| 9 | 172,653 | 174,433 | 0.79 | 0.608 | 0.97 |

**Table 2:**
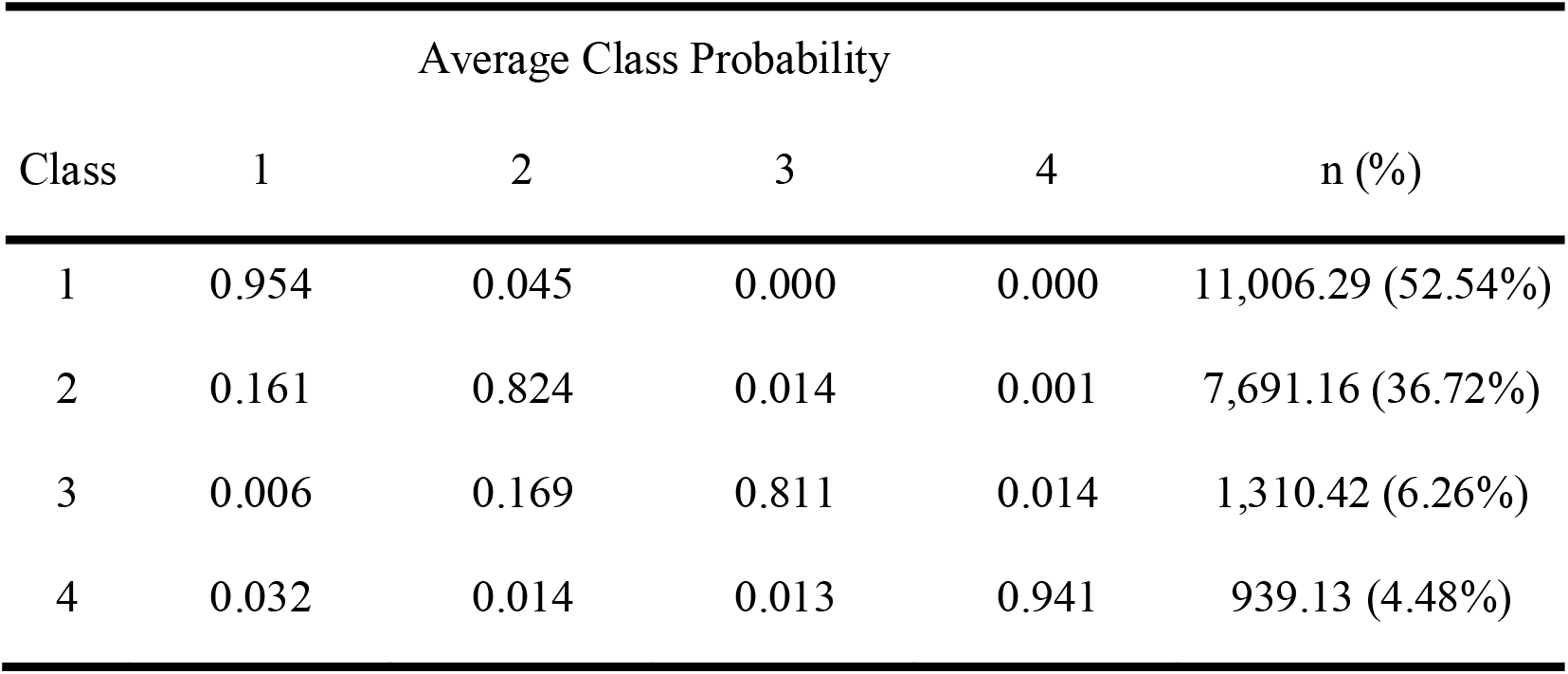
Average latent class membership probability (column) by most likely latent class (row), and expected sample sizes by latent class.

| Class | Average Class Probability |  |  |  | n (%) |
| --- | --- | --- | --- | --- | --- |
|  | 1 | 2 | 3 | 4 |  |
| 1 | 0.954 | 0.045 | 0.000 | 0.000 | 11,006.29 (52.54%) |
| 2 | 0.161 | 0.824 | 0.014 | 0.001 | 7,691.16 (36.72%) |
| 3 | 0.006 | 0.169 | 0.811 | 0.014 | 1,310.42 (6.26%) |
| 4 | 0.032 | 0.014 | 0.013 | 0.941 | 939.13 (4.48%) |

**Figure 2:**
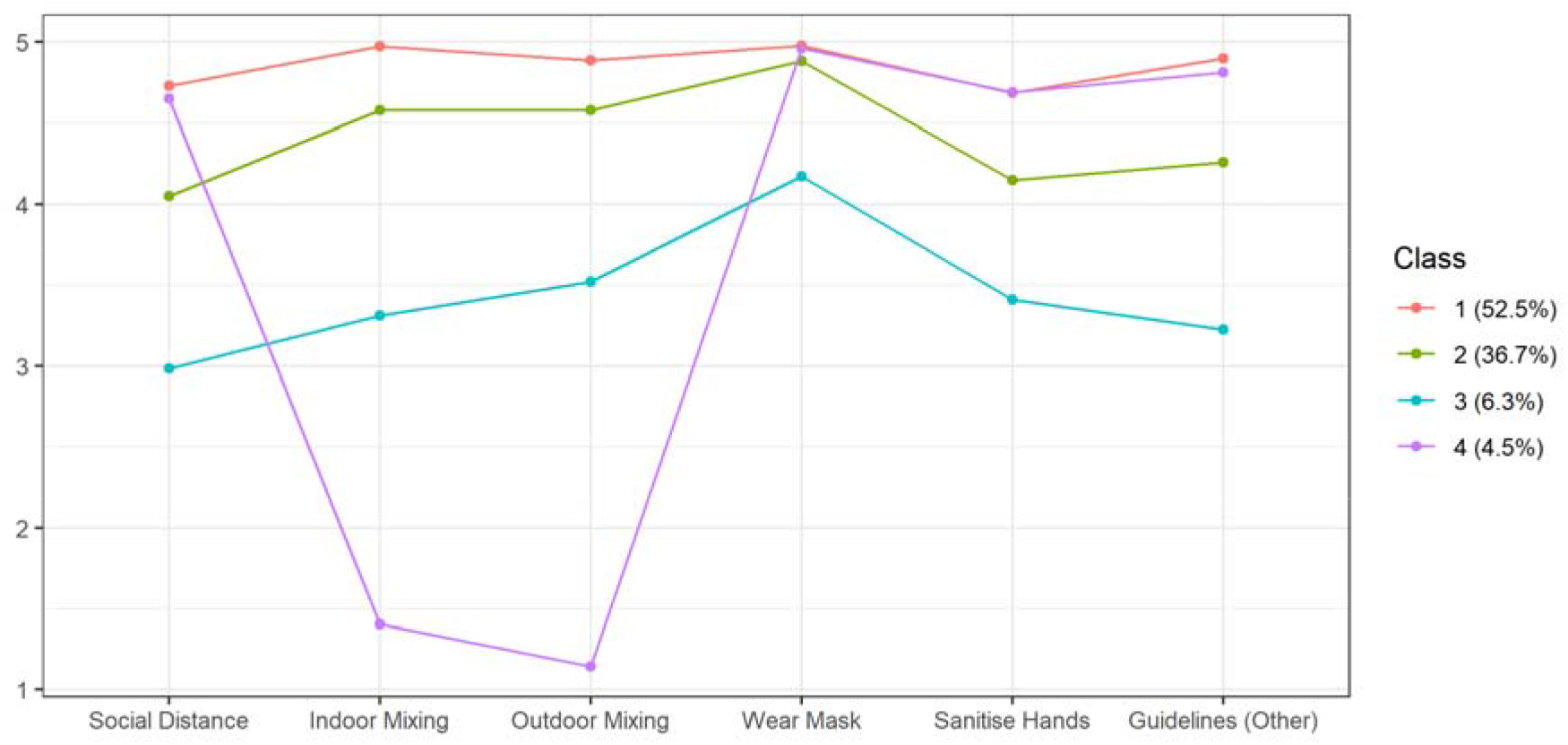
Average compliance levels by latent class

The majority of participants were predicted to be *full compliers* (Class 1: 52.5%), reporting high compliance levels with each of the behaviours. A large minority of individuals (Class 2: 36.7%) were *frequent compliers*, exhibiting frequent or complete compliance with each behaviour. Compared with *full compliers*, these individuals reported similar compliance with mask wearing and somewhat lower compliance with social distancing, hand sanitising and compliance with other guidelines. A small minority were *occasional compliers* (Class 3: 6.3%), reporting occasional or frequent compliance with each behaviour. Again, these individuals reported highest compliance with mask wearing and lower compliance with social distancing and other behaviours. *Full, frequent*, and *occasional compliers* displayed broadly consistent levels of compliance across behaviours, while the final group, *household mixers* (Class 4: 4.5%), instead reported high compliance with each behaviour, except household mixing, for which they reported non-compliance. Note, however, the items on household mixing were reverse coded, and so this pattern may be explained by participant inattention.

### Predictors of Compliance Patterns

Descriptive statistics by most likely class are displayed in Supplementary Table S2. The predictors of the compliance patterns drawn from the 3-step multinomial regression model are displayed in Figure 3 (see Supplementary Table S3 for specific values). Compared with *full compliers*, there was clear evidence that *frequent compliers* (left panel) have lower confidence in government, lower empathy, and a greater external loci of control, are younger, more risk-taking, have lower trait openness, are less conscientious and are less likely to have been self-isolating at first data collection or have a long-term condition. Of these factors, age and risk-taking behaviour were particularly strongly related to *frequent compliance*. Qualitatively similar results were observed comparing *occasional compliers* with *full compliers*, with the exception that there was also stronger evidence that individuals whose mental health did not stay the same during the first national lockdown were more likely to be *occasional compliers*, though both improved and worsened mental health were related to *occasional* compliance. Few factors were clearly related to *household mixing* (exceptions were ethnicity, risk behaviour, and gender), which again may suggest this category reflects inattentive responses.

**Figure 3:**
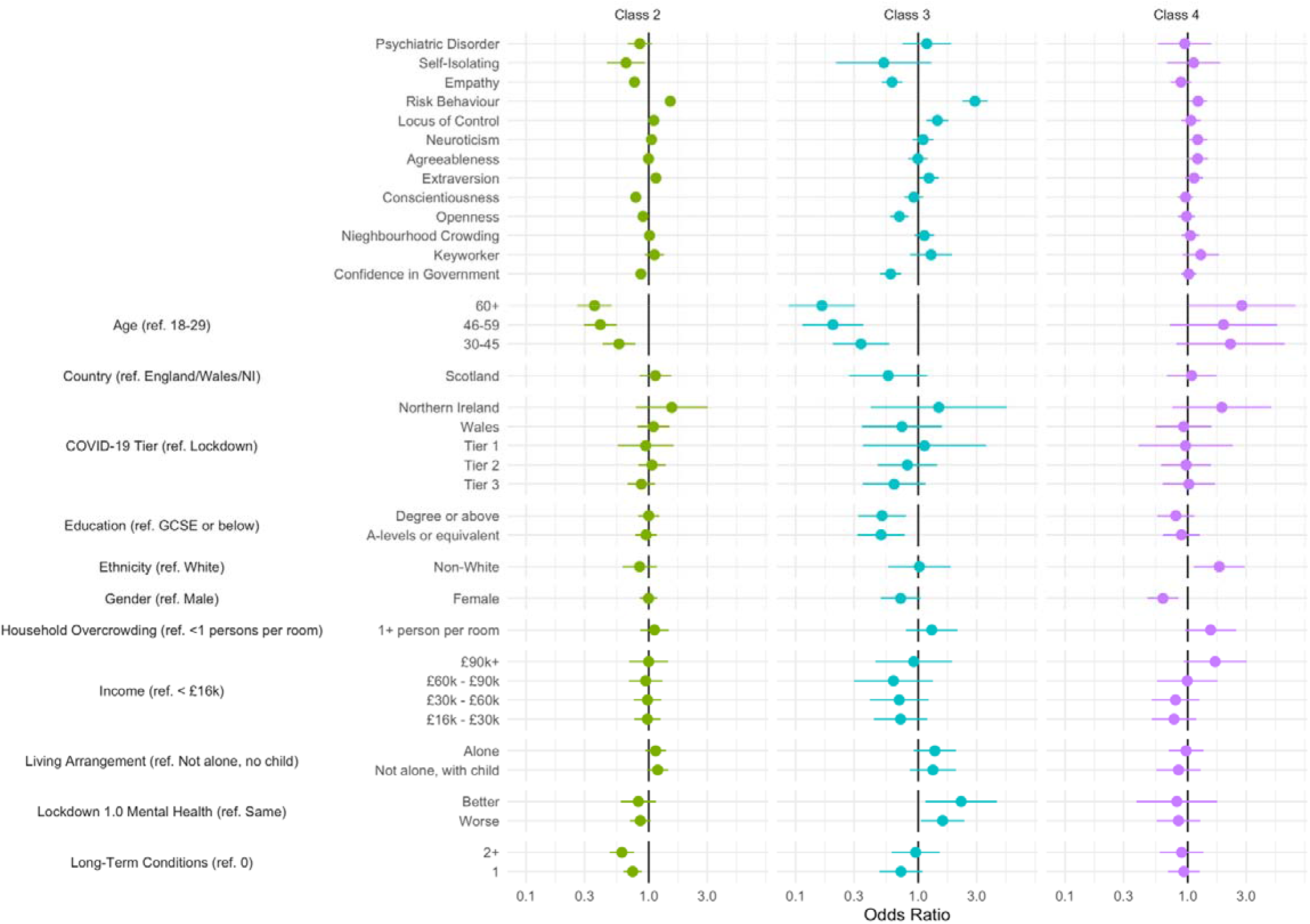
Results of multinomial regression model of latent class on participant characteristics. Reference class: Class 1. Regressions 3-step procedure and use multiply imputed data (40 datasets)

### Predictors of Individual Compliance Behaviours

The results of ordinal regression models predicting individual compliance behaviours are displayed in Supplementary Figure S5 (see Supplementary Tables S4 for specific values). Associations between compliance and individual characteristics were generally consistent across different compliance behaviours. Again, risk behaviour and age were strong predictors of compliance. There were some specificities in the findings, however. For instance, trait extraversion most strongly related to lower compliance with indoor and outdoor household mixing guidelines.

## Discussion

Using latent class analysis, we identified four groups of compliance behaviours. While some behaviours, such as face mask wearing, were performed more frequently overall, most individuals reported broadly consistent levels of compliance across the six behaviours. Further, high levels of compliance across each behaviour was the modal response. A small minority of individuals reported low compliance levels on household mixing but not other measures, though this may be explained by inattentive responses. Behaviour patterns and compliance with individual behaviours were each related to individual characteristics. Most notably, there was evidence that high compliance was strongly related to older age and to lower risk-taking behaviour, consistent with previous research using the COVID-19 Social Study (Wright & Fancourt, 2020). Both of these represent motivations for compliance, and are consistent with a previous study applying the COM-B model to COVID-19 hygiene practices that identified motivation as the most important predictor of compliance behaviour (Miller et al., 2020). However, it should be noted that large differences have been observed between longitudinal and cross-sectional associations with individual characteristics and compliance (Wright et al., Accepted).

The results suggest that individuals choose to comply with all guidelines, rather than some but not others. This suggests that strategies to increase compliance should focus on increasing motivations to comply in general, for instance, through campaigns advertising the risks of non-compliance for personal, family, and public health. Reported compliance was high; a notable finding given that data were collected eight months after the first lockdown in the UK and fears at the beginning of the pandemic that extended lockdown may induce “behavioural fatigue” (Harvey, 2020; see also Seiter & Curran, 2021).

Compared with the other behaviours, compliance with social distancing was relatively low. This may reflect the issue that individuals’ capacity to socially distance is constrained by the behaviour of others and the environment (for instance, due to the layout of shops). Given the higher level of compliance with other measures, the results suggest that lower compliance with social distancing is not a matter of low willingness to comply, though it is also possible that non-compliance with social distancing is opportunistic, for instance, when meeting friends. Compliance with mask wearing was particularly high, frequently being carried out even among the low compliance group. This may reflect that mask wearing is legally mandated, involves little personal sacrifice, is easily observed by others (there is evidence that shame and guilt motivate compliance; Nivette et al., 2021), and that clearer cues to action exist in the environment than for other compliance behaviours (West et al., 2020).

We observed some differences with our prior work using data from the COVID-19 Social Study (Wright & Fancourt, 2020). Notably, we did not observe evidence that low compliance was related to higher income or higher education. However, the measure of compliance (single item on compliance with guidelines overall) differed from the present study and may incorporate knowledge of guidelines to a greater extent. Further, the present study used data from the second wave, in which (compared with mid-Summer) overall compliance increased (Ipsos MORI, 2021).

This study had a number of strengths. Unlike several previous studies, we used a large sample and focused on reported compliance behaviour, rather than compliance intentions or a general measure of compliance. The measures we included reflected several important behaviours for reducing transmission of COVID-19 (Chu et al., 2020). We also studied compliance further into the pandemic than has been explored to date. We were able to assess a rich set of compliance predictors, though the associations we tested were cross-sectional (Wright et al., Accepted). The Latent Class Analysis provided a good solution and revealed important insights into compliance behaviour.

However, this study also had several limitations. As noted, we used cross-sectional data, so results may be biased by unobserved confounding. Our measures of compliance also relied upon self-report data. While participants provided anonymous responses, responses may still be subject to social desirability concerns. Further, though the pandemic is a salient event, the opportunities for non-compliance are many. Individuals may not recall compliance accurately. Guidelines also changed over the study period, which may have influenced responses. Last, we used data from a non-representative (albeit heterogeneous and weighted) sample with substantial attrition rates. High compliers with COVID-19 are likely to be overrepresented in the data, though high levels of compliance have been found in other samples (Ipsos MORI, 2021).

Overall, our results suggest that while compliance with some behaviours is higher in general, individuals comply consistently across recommended compliance behaviours. In line with previous studies (Miller et al., 2020; Wright & Fancourt, 2020), this suggests that motivation is an particularly important determinant of compliance behaviour. Interventions that increase or maintain motivation to comply may be particularly effective at reducing transmission of COVID-19.

## Data Availability

Data used in this study will be made publicly available once the pandemic is over. The code used to run the analysis is available at https://osf.io/aswqc/.

https://github.com/UCL-BSH/CSSUserGuide

## Acknowledgements

The researchers are grateful for the support of a number of organisations with their recruitment efforts including: the UKRI Mental Health Networks, Find Out Now, UCL BioResource, HealthWise Wales, SEO Works, FieldworkHub, and Optimal Workshop.

## Author contributions

LW, AS and DF conceived and designed the study. LW analysed the data and wrote the first draft. All authors provided critical revisions. All authors read and approved the submitted manuscript.

