## Supplementary Information for "Patterns of compliance with COVID-19 preventive behaviours: a latent class analysis of 20,000 UK adults"

### Policy Environment

The period 17 November to 23 December 2020 overlaps with the second wave of COVID-19 in the UK in which there were several changes to COVID-19 related rules. The rules differed somewhat between the nations of England, Scotland, Wales, and Northern Ireland, though mask wearing in indoor public spaces was legally mandated in each nation. On 17 November, England was two weeks into a four-week national lockdown in which non-essential business were closed, employees were asked to work from home where possible, and household mixing was banned. A new three-tier system was put in place from 2 December when lockdown ended. Several areas were placed in the highest tier. In Wales, rules were more relaxed, following a three week “circuit breaker” lockdown that ended on 9 November. Two households were allowed to form bubbles, non-essential shops were open and groups of four people were allowed to meet in indoor public spaces. In Scotland, a five-tier localised system was in place. On 20 November, eleven council areas were placed in tier 4 and non-essential travel between Scotland and England was banned. Northern Ireland was in a circuit breaker lockdown, due to expire on 27 November. Following this, strict rules are put in place, with non-essential shop barred from opening and limited household mixing allowed.

On 24 November, the leads of the four national Governments announced plans to allow mixing of three households for five days between 23-27 December (22-28 December in Northern Ireland), though these plans were later reduced and restricted in some areas. On 2 December, the UK approved the Pfizer/BioNTech COVID-19 vaccine with the first vaccination provided on 8 December. 137,987 people were vaccinated in the first week. On 14 December, a new, more virulent variant of SARS-CoV-2 is identified. Areas in the South East of England are placed into lockdown from 19 December, with Christmas bubbles cancelled in these areas and relaxation elsewhere in England limited to Christmas day. Supplementary Figure S1 shows 7-day COVID-19 caseloads and confirmed deaths, along with the Oxford Policy Tracker, a numerical summary of policy stringency (Hale et al., 2020), across the study period.

### Measures

#### Compliance Behaviour

Compliance behaviour was measured with the following six question:

In the last 7 days, to what extent have you been following the behaviours below?

1. Washing your hands thoroughly with soap and water or using hand sanitising gel after any possible contact with other people outside of your household or shared surfaces
2. Wearing a face mask or other face covering where it is currently recommended
3. Maintaining the recommended distance from people not in your household/bubble
4. Meeting up with MORE THAN the recommended number of people from other households OUTDOORS
5. Meeting up with MORE THAN the recommended number of people from other households INDOORS
6. Following other rules relevant to the tier or level of lockdown currently active in your area

The response categories were: never, rarely, occasionally, frequently, always, not applicable. We treated “not applicable” as a missing value. Items on household mixing were phrased such that higher scores indicated lower compliance. We reverse coded these for consistency with the other behaviours.

#### Predictors of Compliance Behaviour

##### Demographics and Socio-Economic Position

We included the demographic variables for country of residence (England/Wales/NI vs Scotland) and tier (Lockdown, Tier 1, Tier 2, Tier 3, Wales, Northern Ireland), sex (male, female), ethnicity (White or Non-White) and age (18-29, 30-44, 45-59, 60+). We also included variables for socio-economic position (SEP): annual income (< £16k, £16k - £30k, £30k - £60k, £60k - £90k, £90k +), keyworker status (yes, no), education level (GCSE or below, A-levels or equivalent, degree or above), household overcrowding (< 1 person per room, 1+ persons per room), and living arrangement (alone, with adult but no child, with child). Besides tier, which was measure at same data collection as compliance behaviour, each variable was measured at baseline interview.

We also assessed neighbourhood crowding with items collected between 09-16 July. Neighbourhood crowding was measured using three items on satisfaction with neighbourhood traffic density, noise, and levels of crowding. Each was measured on a three-point scale (categories as above), which we reverse coded and summed into a single score (range 3 – 9). Higher values indicate less crowding.

##### Personality Traits

Personality was measured at baseline interview using the Big Five Inventory (BFI-2; Soto & John, 2017), which measures five domains and 15 facets: openness (intellectual curiosity, aesthetic sensitivity, and creative imagination), conscientiousness (organisation, productiveness, and responsibility), extraversion (sociability, assertiveness, and energy level), agreeableness (compassion, respectfulness, and trust) and neuroticism (anxiety, depression, and emotional volatility). Each item was scored on a 5-point scale (1 = “strongly disagree”, 5 = “strongly agree”). We use the sum Likert score for each domain (range 3 - 15). High values indicate high levels of the trait.

Locus of control was measured between 04 – 11 June using the 6-item Locus of Control Scale developed by the University of Washington Beyond High School Project (Hirschman & Almgren, 2012), and captures generalized expectancies about whether individuals can (internal) or cannot (external) control events and outcomes in their lives. Responses were rated on a four-point scale ranging from (1 = “strongly agree”, 4 = “strongly disagree”). We used the sum score of responses, with items coded such that higher values indicate more internal locus of control.

Risk-taking was measured between 23-30 July with one item from the Dohmen Risk Taking Scale (Dohmen et al., 2011). Respondents rated the extent to which they generally see themselves as a person who is fully prepared to take risks, rated on an 11-point scale (0 = “not at all willing to take risks” to 10 = “very willing to take risks”).

Empathy was assessed between 11-18 July using subscales for empathic concern from the Interpersonal Reactivity Index (IRI; Davis, 1983). (Subscales for fantasy and personal distress were not administered.) Empathic concern (also known as emotional empathy) consists of 7 items and captures feelings of warmth, concern, and compassion for others. Items were rated on a five-point scale ranging (1 = “does not describe me well”, 5 = “describes me very well”). We used the sum Likert score, coding items such that higher scores indicate higher empathy (range 0 - 28).

##### Health and Confidence in Government

We included variables for long-term physical health conditions (0, 1, 2+) using a multiple-choice question on medical conditions. Included conditions were high blood pressure, diabetes, heart disease, lung disease, cancer, any other clinically-diagnosed chronic physical health conditions, or any disability. Psyhciatric diagnosis (yes, no) was with the same multiple choice question using items on clinically diagnosed depression, clinically diagnosed anxiety, and any other clinically diagnosed mental health problem. The data was collected at baseline interview.

Mental health during first lockdown vis-à-vis prior to the pandemic was measured with a single item question (“How do you feel your mental health was affected during lockdown in April/May?”). The response categories were: my mental health got worse compared to before Covid-19; My mental health was about the same; my mental health got better compared to before Covid-19. This variable was included in the survey between 18-25 June. Finally, confidence in government was measured with a single item question (“How much confidence do you have in the CENTRAL UK GOVERNMENT that they can handle Covid-19 well?”). Responses were scored on a Likert scale (1. None at all; 7. Lots). This item has been asked at each follow-up. We used responses from the same data collection as compliance behaviours were measured.

### Figures

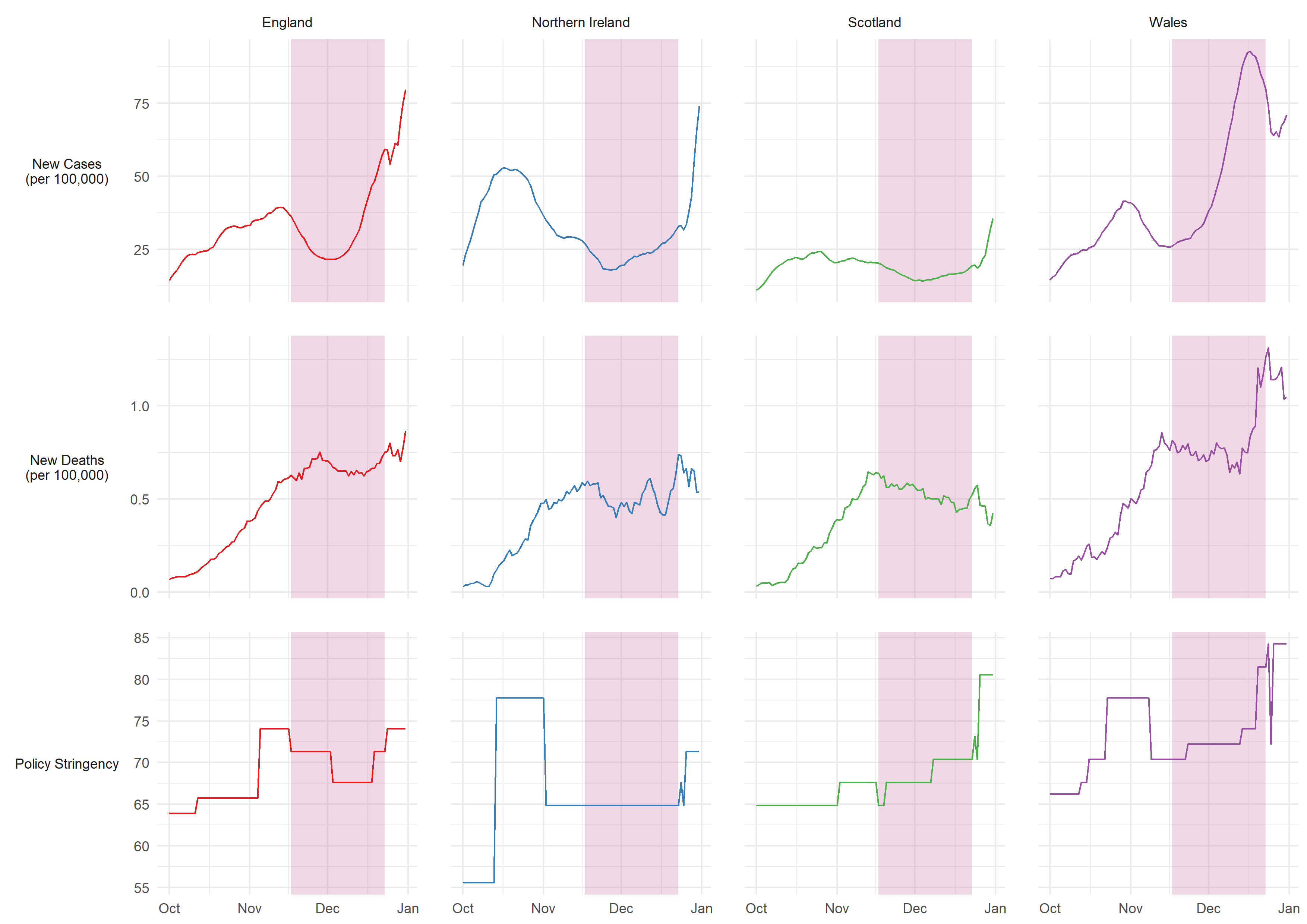

Figure S1: 7-day COVID-19 confirmed cases, COVID-19 deaths, and policy stringency by country. Source: Hale et al. (2020). Pink shaded band represents period in which compliance behaviours were measured.

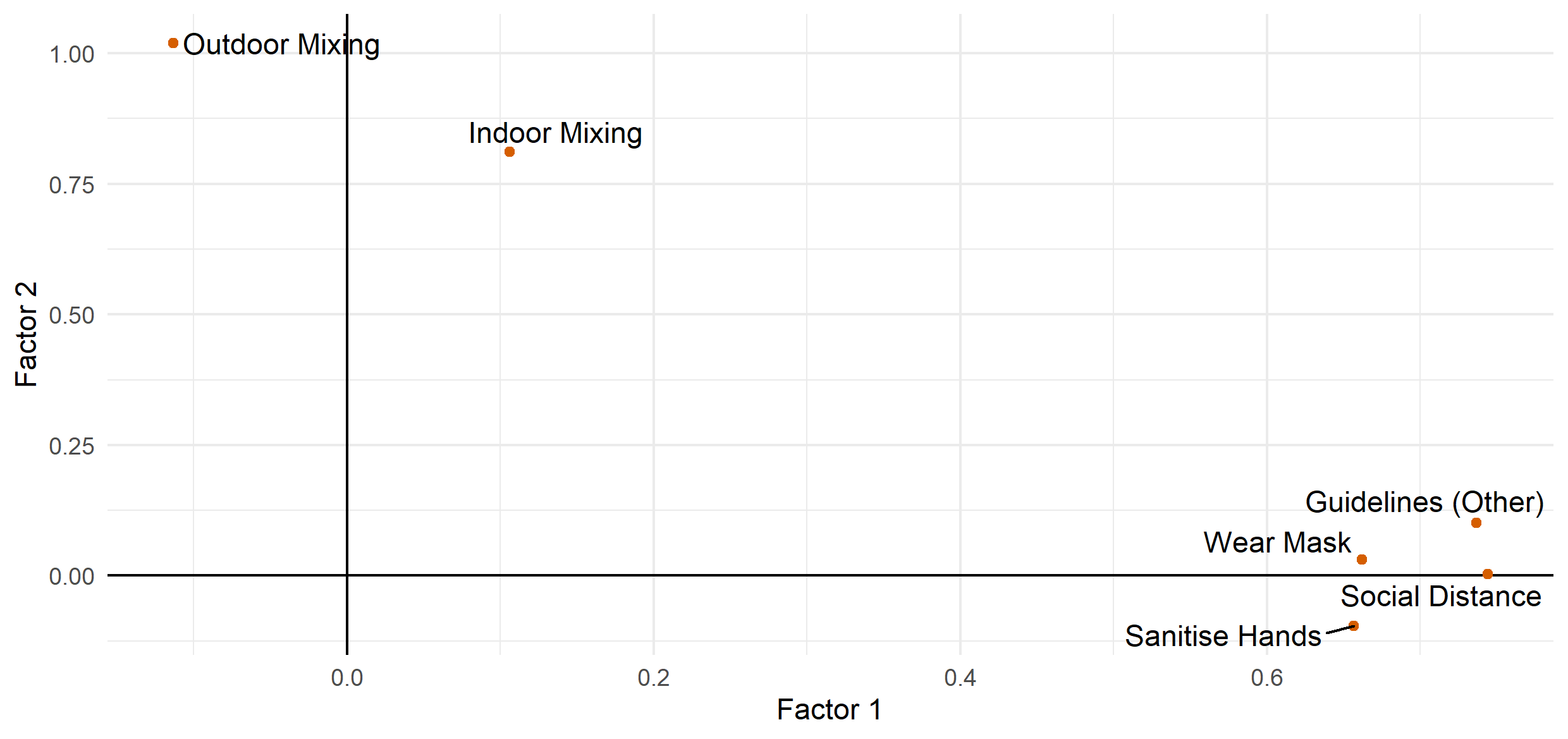

Figure S2: Factor loadings derived from exploratory factor analysis of six compliance items using polychoric correlations and promax rotation.

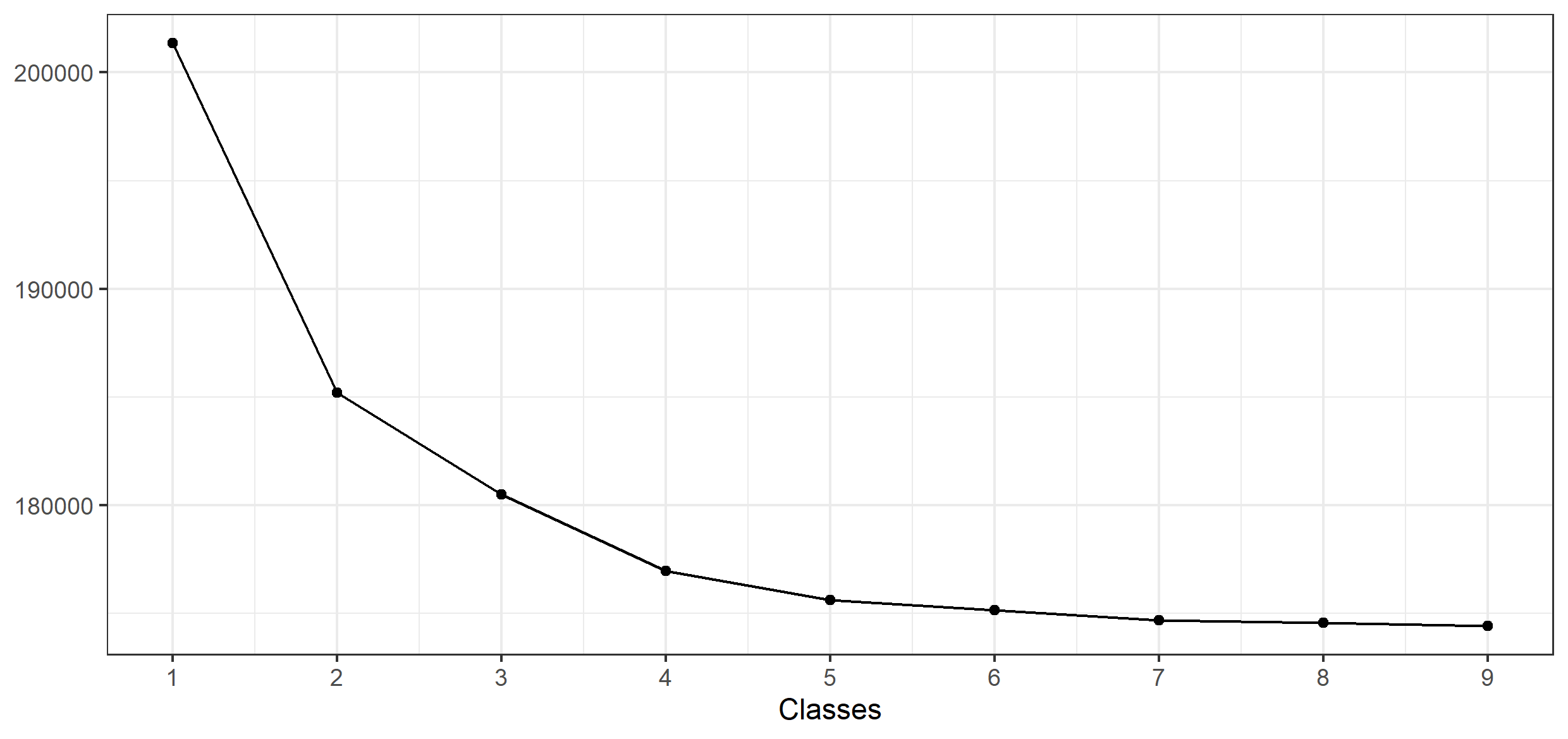

Figure S3: BIC elbow plot for 1-9 latent class solutions.

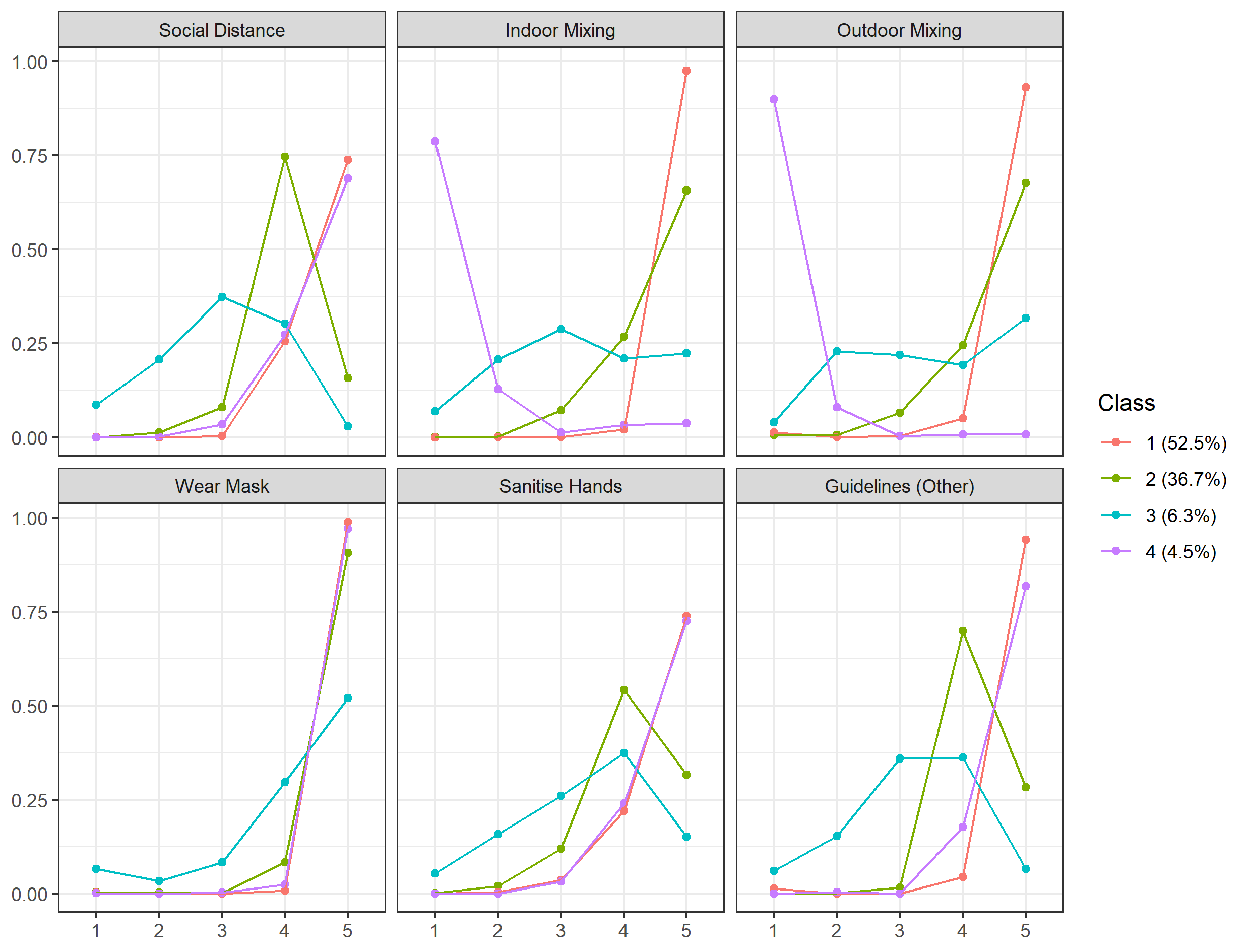

Figure S4: Predicted probability of compliance level by compliance behaviour and latent class

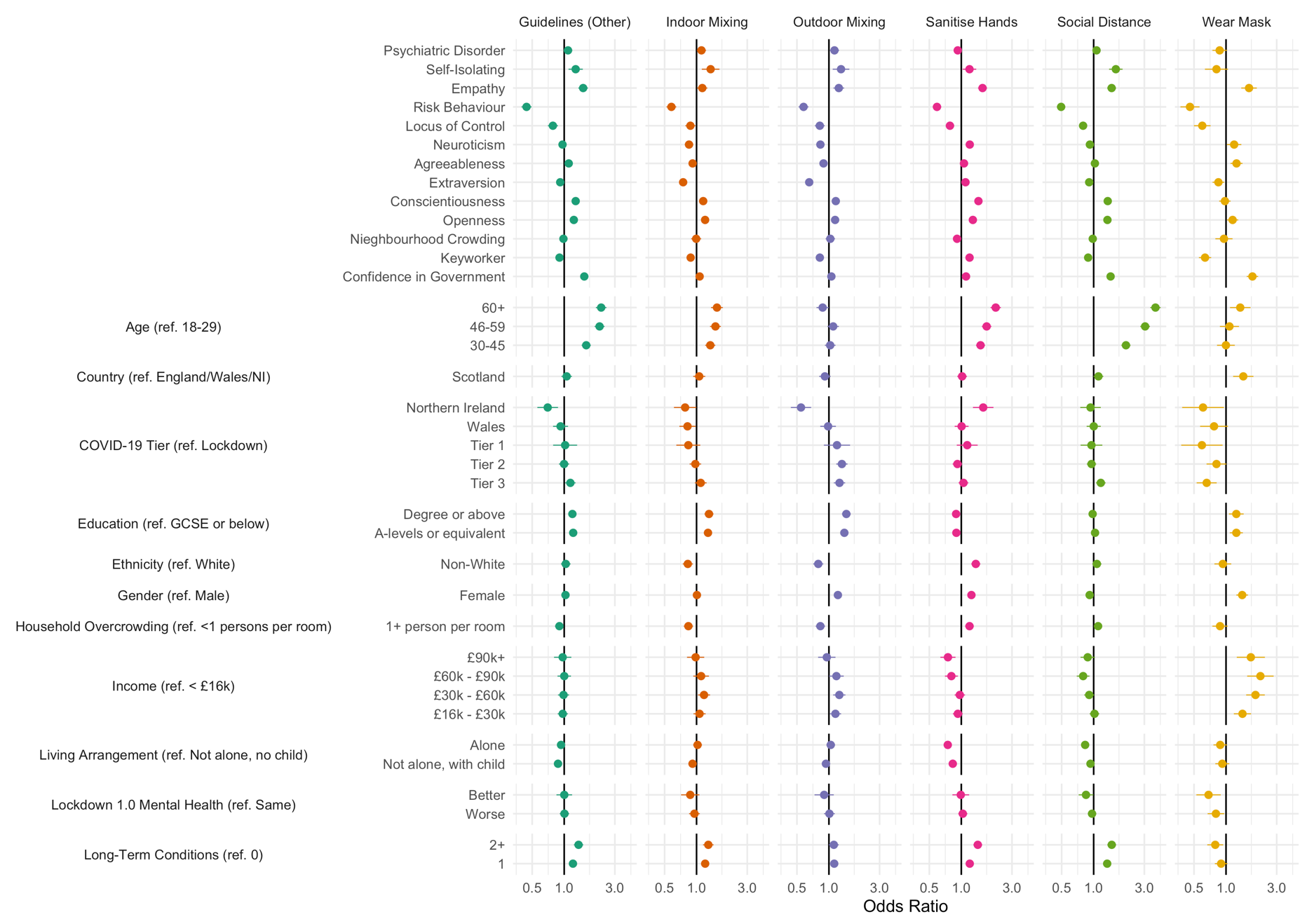

Figure S5: Results of multivariate ordinal regressions predicting compliance behaviour according to participant characteristics. Regressions use multiply imputed data (40 datasets).

### Tables

Table S1: Sample descriptive statistics.

|  | Variable | Unweighted Observed | Missing % | Weighted Imputed |
| --- | --- | --- | --- | --- |
|  | n | 20,947 |  | 20,947 |
| Social Distance | Never | 61 (0.29%) |  | 123.62 (0.59%) |
|  | Rarely | 195 (0.93%) |  | 383.22 (1.83%) |
|  | Occasionally | 793 (3.79%) |  | 1,189.95 (5.68%) |
|  | Frequently | 9,124 (43.56%) |  | 9,217.73 (44%) |
|  | Always | 10,774 (51.43%) |  | 10,032.48 (47.89%) |
| Indoor Mixing | Always | 766 (3.66%) |  | 835.77 (3.99%) |
|  | Frequently | 280 (1.34%) |  | 415.04 (1.98%) |
|  | Occasionally | 798 (3.81%) |  | 956.44 (4.57%) |
|  | Rarely | 2,334 (11.14%) |  | 2,616.46 (12.49%) |
|  | Never | 16,769 (80.05%) |  | 16,123.29 (76.97%) |
| Outdoor Mixing | Always | 990 (4.73%) |  | 1,078.18 (5.15%) |
|  | Frequently | 278 (1.33%) |  | 443.85 (2.12%) |
|  | Occasionally | 760 (3.63%) |  | 837.68 (4%) |
|  | Rarely | 2,411 (11.51%) |  | 2,706.48 (12.92%) |
|  | Never | 16,508 (78.81%) |  | 15,880.80 (75.81%) |
| Wear Mask | Never | 99 (0.47%) |  | 158.83 (0.76%) |
|  | Rarely | 46 (0.22%) |  | 73.00 (0.35%) |
|  | Occasionally | 83 (0.4%) |  | 136.23 (0.65%) |
|  | Frequently | 913 (4.36%) |  | 1,148.91 (5.48%) |
|  | Always | 19,806 (94.55%) |  | 19,430.03 (92.76%) |
| Sanitise Hands | Never | 81 (0.39%) |  | 103.65 (0.49%) |
|  | Rarely | 285 (1.36%) |  | 411.35 (1.96%) |
|  | Occasionally | 1,341 (6.4%) |  | 1,688.16 (8.06%) |
|  | Frequently | 7,138 (34.08%) |  | 7,310.35 (34.9%) |
|  | Always | 12,102 (57.77%) |  | 11,433.49 (54.58%) |
| Other Guidelines | Never | 187 (0.89%) |  | 233.44 (1.11%) |
|  | Rarely | 132 (0.63%) |  | 238.43 (1.14%) |
|  | Occasionally | 392 (1.87%) |  | 597.38 (2.85%) |
|  | Frequently | 5,861 (27.98%) |  | 6,487.19 (30.97%) |
|  | Always | 14,375 (68.63%) |  | 13,390.57 (63.93%) |
|  | Confidence in Government | 3.19 (1.83) |  | 3.29 (1.89) |
| COVID-19 Tier | Lockdown | 3,256 (15.67%) | 0.81% | 3,424.35 (16.35%) |
|  | Tier 3 | 6,019 (28.97%) |  | 6,896.29 (32.92%) |
|  | Tier 2 | 7,985 (38.43%) |  | 8,625.04 (41.18%) |
|  | Tier 1 | 425 (2.05%) |  | 418.82 (2%) |
|  | Wales | 2,892 (13.92%) |  | 1,085.69 (5.18%) |
|  | Northern Ireland | 201 (0.97%) |  | 496.81 (2.37%) |
| Country | England/Wales/NI | 19,679 (93.95%) |  | 19,247.51 (91.89%) |
|  | Scotland | 1,268 (6.05%) |  | 1,699.49 (8.11%) |
| Gender | Male | 5,374 (25.66%) |  | 9,849.22 (47.02%) |
|  | Female | 15,573 (74.34%) |  | 11,097.78 (52.98%) |
| Ethnicity | White | 20,264 (96.74%) |  | 18,834.33 (89.91%) |
|  | Non-White | 683 (3.26%) |  | 2,112.67 (10.09%) |
| Age | 18-29 | 804 (3.84%) |  | 3,169.20 (15.13%) |
|  | 30-45 | 3,970 (18.95%) |  | 5,527.74 (26.39%) |
|  | 46-59 | 6,680 (31.89%) |  | 5,406.59 (25.81%) |
|  | 60+ | 9,493 (45.32%) |  | 6,843.47 (32.67%) |
| Keyworker | No | 16,846 (80.42%) |  | 16,482.43 (78.69%) |
|  | Yes | 4,101 (19.58%) |  | 4,464.57 (21.31%) |
| Education | GCSE or below | 2,993 (14.29%) |  | 6,718.58 (32.07%) |
|  | A-levels or equivalent | 3,531 (16.86%) |  | 6,758.60 (32.27%) |
|  | Degree or above | 14,423 (68.85%) |  | 7,469.81 (35.66%) |
| Income | < £16k | 2,696 (14.34%) | 10.24% | 3,703.64 (17.68%) |
|  | £16k - £30k | 4,972 (26.45%) |  | 6,016.19 (28.72%) |
|  | £30k - £60k | 6,633 (35.28%) |  | 7,140.88 (34.09%) |
|  | £60k - £90k | 2,689 (14.3%) |  | 2,554.19 (12.19%) |
|  | £90k+ | 1,811 (9.63%) |  | 1,532.10 (7.31%) |
| Living Arrangement | Not alone, no child | 12,314 (58.79%) |  | 12,475.06 (59.56%) |
|  | Not alone, with child | 4,120 (19.67%) |  | 4,528.07 (21.62%) |
|  | Alone | 4,513 (21.54%) |  | 3,943.87 (18.83%) |
| Household Overcrowding | <1 persons per room | 19,597 (93.56%) |  | 18,319.81 (87.46%) |
|  | 1+ person per room | 1,350 (6.44%) |  | 2,627.19 (12.54%) |
|  | Nieghbourhood Crowding | 7.03 (1.84) | 24.48% | 6.95 (1.85) |
| Lockdown 1.0 Mental Health | Same | 10,249 (62.2%) | 21.33% | 12,448.18 (59.43%) |
|  | Worse | 5,001 (30.35%) |  | 6,858.43 (32.74%) |
|  | Better | 1,228 (7.45%) |  | 1,640.39 (7.83%) |
|  | Openness | 15.3 (3.23) |  | 14.75 (3.26) |
|  | Conscientiousness | 16.08 (2.88) |  | 15.78 (2.9) |
|  | Extraversion | 12.88 (4.25) |  | 12.51 (4.28) |
|  | Agreeableness | 15.59 (3.01) |  | 15.44 (3.13) |
|  | Neuroticism | 10.9 (4.21) |  | 11.27 (4.37) |
|  | Locus of Control | 12.14 (2.58) | 20.82% | 12.53 (2.67) |
|  | Risk Behaviour | 4.38 (2.33) | 25.4% | 4.42 (2.36) |
|  | Empathy | 20.67 (4.6) | 22.33% | 20 (4.79) |
| Self-Isolating | No | 19,010 (93.4%) | 2.84% | 19,640.32 (93.76%) |
|  | Yes | 1,343 (6.6%) |  | 1,306.68 (6.24%) |
| Psychiatric Disorder | No | 17,997 (85.92%) |  | 17,407.99 (83.1%) |
|  | Yes | 2,950 (14.08%) |  | 3,539.01 (16.9%) |
| Long-Term Conditions | 0 | 11,935 (56.98%) |  | 12,247.76 (58.47%) |
|  | 1 | 5,852 (27.94%) |  | 5,579.65 (26.64%) |
|  | 2+ | 3,160 (15.09%) |  | 3,119.59 (14.89%) |

Table S2: Sample descriptive statistics by most likely latent class.

|  | Variable | Class 1 | Class 2 | Class 3 | Class 4 |
| --- | --- | --- | --- | --- | --- |
|  | n | 11,780.40 | 7,063.10 | 1,187.06 | 916.44 |
| Social Distance | Never | 9.45 (0.08%) | - | 114.18 (9.62%) | - |
|  | Rarely | - | 110.50 (1.56%) | 270.31 (22.77%) | 2.41 (0.26%) |
|  | Occasionally | 47.94 (0.41%) | 647.46 (9.17%) | 462.82 (38.99%) | 31.73 (3.46%) |
|  | Frequently | 3,450.72 (29.29%) | 5,197.75 (73.59%) | 311.22 (26.22%) | 258.04 (28.16%) |
|  | Always | 8,272.30 (70.22%) | 1,107.39 (15.68%) | 28.53 (2.4%) | 624.26 (68.12%) |
| Indoor Mixing | Always | - | 6.78 (0.1%) | 89.51 (7.54%) | 739.48 (80.69%) |
|  | Frequently | 9.56 (0.08%) | 7.54 (0.11%) | 272.27 (22.94%) | 125.67 (13.71%) |
|  | Occasionally | 9.46 (0.08%) | 590.38 (8.36%) | 345.10 (29.07%) | 11.49 (1.25%) |
|  | Rarely | 148.98 (1.26%) | 2,194.64 (31.07%) | 235.42 (19.83%) | 37.43 (4.08%) |
|  | Never | 11,612.40 (98.57%) | 4,263.76 (60.37%) | 244.76 (20.62%) | 2.36 (0.26%) |
| Outdoor Mixing | Always | 168.02 (1.43%) | 33.51 (0.47%) | 45.23 (3.81%) | 831.42 (90.72%) |
|  | Frequently | 11.54 (0.1%) | 62.69 (0.89%) | 298.22 (25.12%) | 71.39 (7.79%) |
|  | Occasionally | 29.78 (0.25%) | 546.53 (7.74%) | 258.43 (21.77%) | 2.94 (0.32%) |
|  | Rarely | 539.32 (4.58%) | 1,930.61 (27.33%) | 230.64 (19.43%) | 5.92 (0.65%) |
|  | Never | 11,031.73 (93.64%) | 4,489.77 (63.57%) | 354.54 (29.87%) | 4.76 (0.52%) |
| Wear Mask | Never | 30.14 (0.26%) | 39.91 (0.57%) | 87.16 (7.34%) | 1.62 (0.18%) |
|  | Rarely | 4.31 (0.04%) | 23.49 (0.33%) | 45.20 (3.81%) | - |
|  | Occasionally | 9.53 (0.08%) | 13.87 (0.2%) | 109.67 (9.24%) | 3.16 (0.34%) |
|  | Frequently | 84.96 (0.72%) | 665.94 (9.43%) | 374.12 (31.52%) | 23.89 (2.61%) |
|  | Always | 11,651.46 (98.91%) | 6,319.89 (89.48%) | 570.91 (48.09%) | 887.76 (96.87%) |
| Sanitise Hands | Never | 17.30 (0.15%) | 13.70 (0.19%) | 72.65 (6.12%) | - |
|  | Rarely | 23.09 (0.2%) | 186.21 (2.64%) | 201.20 (16.95%) | 0.86 (0.09%) |
|  | Occasionally | 507.95 (4.31%) | 839.69 (11.89%) | 305.29 (25.72%) | 35.22 (3.84%) |
|  | Frequently | 2,882.83 (24.47%) | 3,761.64 (53.26%) | 439.40 (37.02%) | 226.47 (24.71%) |
|  | Always | 8,349.23 (70.87%) | 2,261.87 (32.02%) | 168.51 (14.2%) | 653.88 (71.35%) |
| Other Guidelines | Never | 153.87 (1.31%) | - | 79.57 (6.7%) | - |
|  | Rarely | 18.43 (0.16%) | 10.87 (0.15%) | 209.13 (17.62%) | - |
|  | Occasionally | - | 110.53 (1.56%) | 486.65 (41%) | 0.20 (0.02%) |
|  | Frequently | 314.56 (2.67%) | 5,654.68 (80.06%) | 352.93 (29.73%) | 165.01 (18.01%) |
|  | Always | 11,293.54 (95.87%) | 1,287.03 (18.22%) | 58.77 (4.95%) | 751.23 (81.97%) |
|  | Confidence in Government | 3.42 (1.95) | 3.15 (1.76) | 2.65 (1.75) | 3.56 (1.96) |
| COVID-19 Tier | Lockdown | 1,866.94 (15.85%) | 1,219.93 (17.27%) | 208.98 (17.6%) | 128.51 (14.02%) |
|  | Tier 3 | 4,033.94 (34.24%) | 2,193.97 (31.06%) | 349.65 (29.46%) | 318.72 (34.78%) |
|  | Tier 2 | 4,739.27 (40.23%) | 2,994.06 (42.39%) | 523.12 (44.07%) | 368.60 (40.22%) |
|  | Tier 1 | 250.89 (2.13%) | 124.77 (1.77%) | 22.91 (1.93%) | 20.25 (2.21%) |
|  | Wales | 639.99 (5.43%) | 347.95 (4.93%) | 48.93 (4.12%) | 48.81 (5.33%) |
|  | Northern Ireland | 249.37 (2.12%) | 182.43 (2.58%) | 33.47 (2.82%) | 31.54 (3.44%) |
| Country | England/Wales/NI | 10,849.03 (92.09%) | 6,440.25 (91.18%) | 1,115.12 (93.94%) | 843.11 (92%) |
|  | Scotland | 931.37 (7.91%) | 622.85 (8.82%) | 71.94 (6.06%) | 73.32 (8%) |
| Gender | Male | 5,344.62 (45.37%) | 3,349.11 (47.42%) | 626.77 (52.8%) | 528.72 (57.69%) |
|  | Female | 6,435.78 (54.63%) | 3,713.99 (52.58%) | 560.29 (47.2%) | 387.72 (42.31%) |
| Ethnicity | White | 10,654.43 (90.44%) | 6,349.89 (89.9%) | 1,034.94 (87.19%) | 795.07 (86.76%) |
|  | Non-White | 1,125.97 (9.56%) | 713.22 (10.1%) | 152.12 (12.81%) | 121.36 (13.24%) |
| Age | 18-29 | 1,334.02 (11.32%) | 1,440.94 (20.4%) | 341.84 (28.8%) | 52.40 (5.72%) |
|  | 30-45 | 2,862.69 (24.3%) | 2,078.89 (29.43%) | 363.15 (30.59%) | 223.01 (24.33%) |
|  | 46-59 | 3,284.26 (27.88%) | 1,647.23 (23.32%) | 244.55 (20.6%) | 230.54 (25.16%) |
|  | 60+ | 4,299.43 (36.5%) | 1,896.04 (26.84%) | 237.52 (20.01%) | 410.48 (44.79%) |
| Keyworker | No | 9,461.27 (80.31%) | 5,432.78 (76.92%) | 871.02 (73.38%) | 717.36 (78.28%) |
|  | Yes | 2,319.13 (19.69%) | 1,630.32 (23.08%) | 316.04 (26.62%) | 199.08 (21.72%) |
| Education | GCSE or below | 3,970.63 (33.71%) | 1,984.39 (28.1%) | 400.81 (33.76%) | 362.76 (39.58%) |
|  | A-levels or equivalent | 3,825.68 (32.47%) | 2,298.73 (32.55%) | 348.59 (29.37%) | 285.61 (31.16%) |
|  | Degree or above | 3,984.09 (33.82%) | 2,779.99 (39.36%) | 437.66 (36.87%) | 268.07 (29.25%) |
| Income | < £16k | 2,092.37 (17.76%) | 1,153.56 (16.33%) | 264.06 (22.25%) | 193.64 (21.13%) |
|  | £16k - £30k | 3,529.09 (29.96%) | 1,922.61 (27.22%) | 308.45 (25.98%) | 256.04 (27.94%) |
|  | £30k - £60k | 4,019.96 (34.12%) | 2,477.60 (35.08%) | 373.06 (31.43%) | 270.25 (29.49%) |
|  | £60k - £90k | 1,361.05 (11.55%) | 952.11 (13.48%) | 139.49 (11.75%) | 101.54 (11.08%) |
|  | £90k+ | 777.93 (6.6%) | 557.22 (7.89%) | 101.98 (8.59%) | 94.96 (10.36%) |
| Living Arrangement | Not alone, no child | 7,192.11 (61.05%) | 4,062.25 (57.51%) | 644.92 (54.33%) | 575.78 (62.83%) |
|  | Not alone, with child | 2,385.20 (20.25%) | 1,687.05 (23.89%) | 287.08 (24.18%) | 168.74 (18.41%) |
|  | Alone | 2,203.09 (18.7%) | 1,313.81 (18.6%) | 255.06 (21.49%) | 171.92 (18.76%) |
| Household Overcrowding | <1 persons per room | 10,512.03 (89.23%) | 6,047.53 (85.62%) | 970.34 (81.74%) | 789.91 (86.19%) |
|  | 1+ person per room | 1,268.37 (10.77%) | 1,015.57 (14.38%) | 216.72 (18.26%) | 126.53 (13.81%) |
|  | Nieghbourhood Crowding | 6.95 (1.87) | 6.95 (1.82) | 6.91 (1.8) | 7.03 (1.87) |
| Lockdown 1.0 Mental Health | Same | 7,168.37 (60.85%) | 4,153.77 (58.81%) | 510.84 (43.03%) | 615.20 (67.13%) |
|  | Worse | 3,748.86 (31.82%) | 2,340.89 (33.14%) | 518.38 (43.67%) | 250.30 (27.31%) |
|  | Better | 863.17 (7.33%) | 568.44 (8.05%) | 157.84 (13.3%) | 50.94 (5.56%) |
|  | Openness | 14.86 (3.31) | 14.65 (3.1) | 14.2 (3.42) | 14.79 (3.5) |
|  | Conscientiousness | 16.04 (2.9) | 15.39 (2.79) | 15.29 (3.07) | 15.99 (3.05) |
|  | Extraversion | 12.46 (4.28) | 12.57 (4.22) | 12.38 (4.6) | 12.84 (4.29) |
|  | Agreeableness | 15.63 (3.13) | 15.18 (3.01) | 14.87 (3.35) | 15.77 (3.41) |
|  | Neuroticism | 11.27 (4.4) | 11.24 (4.31) | 11.54 (4.45) | 11.11 (4.37) |
|  | Locus of Control | 12.43 (2.68) | 12.59 (2.59) | 13.24 (2.99) | 12.53 (2.64) |
|  | Risk Behaviour | 4.04 (2.35) | 4.81 (2.21) | 5.74 (2.48) | 4.45 (2.41) |
|  | Empathy | 20.49 (4.76) | 19.43 (4.7) | 18.54 (4.99) | 20.02 (4.8) |
| Self-Isolating | No | 10,863.39 (92.22%) | 6,788.61 (96.11%) | 1,149.35 (96.82%) | 838.96 (91.55%) |
|  | Yes | 917.01 (7.78%) | 274.49 (3.89%) | 37.71 (3.18%) | 77.47 (8.45%) |
| Psychiatric Disorder | No | 9,760.18 (82.85%) | 5,959.08 (84.37%) | 912.82 (76.9%) | 775.92 (84.67%) |
|  | Yes | 2,020.22 (17.15%) | 1,104.03 (15.63%) | 274.24 (23.1%) | 140.52 (15.33%) |
| Long-Term Conditions | 0 | 6,324.24 (53.68%) | 4,667.52 (66.08%) | 780.92 (65.79%) | 475.09 (51.84%) |
|  | 1 | 3,413.22 (28.97%) | 1,648.78 (23.34%) | 249.66 (21.03%) | 267.99 (29.24%) |
|  | 2+ | 2,042.94 (17.34%) | 746.81 (10.57%) | 156.48 (13.18%) | 173.36 (18.92%) |

Table S3: Results of multinomial regression model predicting latent class by individual characteristics. Models use 3-step procedure and multiply imputed data (40 datasets). Reference class is largest class, Class 1.

|  | Variable | Class 2 | Class 3 | Class 4 |
| --- | --- | --- | --- | --- |
|  | Confidence in Government | 0.86 (0.8, 0.93) | 0.6 (0.49, 0.73) | 1.02 (0.89, 1.17) |
| COVID-19 Tier (ref. Lockdown) | Tier 3 | 0.87 (0.67, 1.13) | 0.64 (0.35, 1.15) | 1.02 (0.62, 1.67) |
|  | Tier 2 | 1.06 (0.82, 1.38) | 0.82 (0.47, 1.43) | 0.97 (0.61, 1.55) |
|  | Tier 1 | 0.94 (0.56, 1.6) | 1.12 (0.35, 3.58) | 0.96 (0.4, 2.32) |
|  | Wales | 1.09 (0.81, 1.47) | 0.74 (0.35, 1.56) | 0.93 (0.55, 1.56) |
|  | Northern Ireland | 1.54 (0.78, 3.03) | 1.47 (0.41, 5.28) | 1.9 (0.75, 4.79) |
| Country (ref. England/Wales/NI) | Scotland | 1.13 (0.84, 1.53) | 0.57 (0.27, 1.19) | 1.08 (0.68, 1.71) |
| Gender (ref. Male) | Female | 1 (0.84, 1.17) | 0.72 (0.49, 1.04) | 0.63 (0.47, 0.84) |
| Ethnicity (ref. White) | Non-White | 0.84 (0.61, 1.16) | 1.02 (0.57, 1.83) | 1.81 (1.12, 2.92) |
| Age (ref. 18-29) | 30-45 | 0.57 (0.42, 0.78) | 0.34 (0.2, 0.58) | 2.23 (0.81, 6.17) |
|  | 46-59 | 0.4 (0.3, 0.55) | 0.2 (0.11, 0.36) | 1.96 (0.71, 5.35) |
|  | 60+ | 0.36 (0.26, 0.5) | 0.16 (0.09, 0.31) | 2.76 (1.01, 7.55) |
|  | Keyworker | 1.11 (0.93, 1.34) | 1.27 (0.86, 1.89) | 1.28 (0.91, 1.8) |
| Education (ref. GCSE or below) | A-levels or equivalent | 0.95 (0.78, 1.16) | 0.5 (0.32, 0.78) | 0.89 (0.63, 1.26) |
|  | Degree or above | 1 (0.82, 1.22) | 0.51 (0.32, 0.79) | 0.8 (0.56, 1.13) |
| Income (ref. < £16k) | £16k - £30k | 0.98 (0.76, 1.25) | 0.72 (0.43, 1.19) | 0.77 (0.51, 1.17) |
|  | £30k - £60k | 0.98 (0.75, 1.27) | 0.7 (0.4, 1.22) | 0.79 (0.51, 1.24) |
|  | £60k - £90k | 0.94 (0.69, 1.29) | 0.63 (0.3, 1.32) | 0.99 (0.56, 1.75) |
|  | £90k+ | 1 (0.69, 1.44) | 0.92 (0.45, 1.89) | 1.67 (0.92, 3.03) |
| Living Arrangement (ref. Not alone, no child) | Not alone, with child | 1.18 (0.97, 1.44) | 1.32 (0.85, 2.03) | 0.84 (0.55, 1.27) |
|  | Alone | 1.14 (0.94, 1.38) | 1.37 (0.92, 2.03) | 0.97 (0.7, 1.34) |
| Household Overcrowding (ref. <1 persons per room) | 1+ person per room | 1.11 (0.85, 1.46) | 1.29 (0.79, 2.09) | 1.54 (0.95, 2.48) |
|  | Nieghbourhood Crowding | 1.02 (0.93, 1.11) | 1.12 (0.93, 1.35) | 1.05 (0.89, 1.24) |
| Lockdown 1.0 Mental Health (ref. Same) | Worse | 0.85 (0.7, 1.04) | 1.58 (1.05, 2.38) | 0.84 (0.56, 1.27) |
|  | Better | 0.82 (0.59, 1.15) | 2.24 (1.14, 4.38) | 0.81 (0.38, 1.74) |
|  | Openness | 0.9 (0.83, 0.97) | 0.7 (0.59, 0.83) | 0.98 (0.83, 1.14) |
|  | Conscientiousness | 0.78 (0.73, 0.85) | 0.92 (0.77, 1.1) | 0.96 (0.83, 1.11) |
|  | Extraversion | 1.14 (1.06, 1.24) | 1.22 (1.02, 1.47) | 1.13 (0.95, 1.33) |
|  | Agreeableness | 1 (0.91, 1.08) | 0.99 (0.83, 1.19) | 1.2 (0.99, 1.46) |
|  | Neuroticism | 1.05 (0.96, 1.16) | 1.1 (0.9, 1.34) | 1.21 (1.01, 1.45) |
|  | Locus of Control | 1.1 (1, 1.21) | 1.43 (1.17, 1.76) | 1.06 (0.88, 1.28) |
|  | Risk Behaviour | 1.5 (1.37, 1.64) | 2.9 (2.29, 3.68) | 1.21 (1.02, 1.44) |
|  | Empathy | 0.77 (0.69, 0.84) | 0.61 (0.5, 0.74) | 0.88 (0.73, 1.07) |
|  | Self-Isolating | 0.65 (0.46, 0.93) | 0.52 (0.21, 1.28) | 1.12 (0.68, 1.84) |
|  | Psychiatric Disorder | 0.85 (0.67, 1.06) | 1.17 (0.75, 1.85) | 0.95 (0.57, 1.56) |
| Long-Term Conditions (ref. 0) | 1 | 0.74 (0.62, 0.88) | 0.72 (0.48, 1.08) | 0.93 (0.69, 1.25) |
|  | 2+ | 0.6 (0.48, 0.76) | 0.95 (0.61, 1.5) | 0.89 (0.59, 1.34) |

Table S4: Results of ordinal regression models predicting compliance behaviour (column) by individual characteristics. Models use multiply imputed data (40 datasets).

|  | Variable | Social Distance | Indoor Mixing | Outdoor Mixing | Wear Mask | Sanitise Hands | Guidelines (Other) |
| --- | --- | --- | --- | --- | --- | --- | --- |
|  | Confidence in Government | 1.44 (1.36, 1.54) | 1.07 (0.99, 1.15) | 1.05 (0.98, 1.13) | 1.77 (1.57, 2.01) | 1.11 (1.04, 1.18) | 1.55 (1.45, 1.65) |
| COVID-19 Tier (ref. Lockdown) | Tier 3 | 1.17 (1.05, 1.3) | 1.09 (0.96, 1.24) | 1.26 (1.11, 1.42) | 0.66 (0.53, 0.82) | 1.05 (0.94, 1.16) | 1.14 (1.02, 1.28) |
|  | Tier 2 | 0.95 (0.86, 1.06) | 0.97 (0.86, 1.1) | 1.32 (1.17, 1.5) | 0.81 (0.66, 1.01) | 0.92 (0.83, 1.02) | 1 (0.89, 1.11) |
|  | Tier 1 | 0.95 (0.75, 1.21) | 0.84 (0.64, 1.08) | 1.19 (0.9, 1.58) | 0.59 (0.38, 0.93) | 1.14 (0.91, 1.43) | 1.02 (0.79, 1.32) |
|  | Wales | 1 (0.86, 1.17) | 0.82 (0.69, 0.98) | 0.98 (0.83, 1.16) | 0.77 (0.57, 1.04) | 1.01 (0.87, 1.17) | 0.92 (0.79, 1.09) |
|  | Northern Ireland | 0.93 (0.75, 1.17) | 0.78 (0.61, 0.99) | 0.55 (0.44, 0.68) | 0.61 (0.39, 0.95) | 1.61 (1.29, 2.01) | 0.7 (0.56, 0.88) |
| Country (ref. England/Wales/NI) | Scotland | 1.11 (1, 1.23) | 1.06 (0.93, 1.21) | 0.92 (0.81, 1.04) | 1.45 (1.17, 1.81) | 1.02 (0.92, 1.13) | 1.06 (0.94, 1.18) |
| Gender (ref. Male) | Female | 0.92 (0.86, 0.98) | 1.01 (0.93, 1.09) | 1.21 (1.13, 1.31) | 1.42 (1.25, 1.61) | 1.25 (1.17, 1.33) | 1.03 (0.96, 1.1) |
| Ethnicity (ref. White) | Non-White | 1.07 (0.97, 1.18) | 0.83 (0.74, 0.92) | 0.79 (0.71, 0.89) | 0.93 (0.78, 1.12) | 1.37 (1.24, 1.51) | 1.03 (0.93, 1.15) |
| Age (ref. 18-29) | 30-45 | 2.01 (1.83, 2.22) | 1.35 (1.21, 1.5) | 1.03 (0.92, 1.15) | 1 (0.82, 1.21) | 1.52 (1.38, 1.67) | 1.61 (1.46, 1.78) |
|  | 46-59 | 3.05 (2.75, 3.39) | 1.51 (1.34, 1.69) | 1.1 (0.97, 1.24) | 1.08 (0.88, 1.32) | 1.73 (1.56, 1.92) | 2.15 (1.93, 2.4) |
|  | 60+ | 3.82 (3.41, 4.27) | 1.56 (1.37, 1.77) | 0.87 (0.76, 1) | 1.36 (1.09, 1.7) | 2.11 (1.89, 2.36) | 2.23 (1.99, 2.5) |
|  | Keyworker | 0.89 (0.83, 0.95) | 0.88 (0.81, 0.96) | 0.82 (0.76, 0.89) | 0.63 (0.56, 0.73) | 1.2 (1.11, 1.29) | 0.9 (0.84, 0.97) |
| Education (ref. GCSE or below) | A-levels or equivalent | 1.03 (0.96, 1.11) | 1.28 (1.17, 1.4) | 1.4 (1.28, 1.52) | 1.25 (1.09, 1.44) | 0.9 (0.84, 0.97) | 1.22 (1.12, 1.32) |
|  | Degree or above | 0.98 (0.9, 1.06) | 1.31 (1.19, 1.44) | 1.46 (1.33, 1.6) | 1.25 (1.06, 1.47) | 0.89 (0.82, 0.97) | 1.19 (1.09, 1.31) |
| Income (ref. < £16k) | £16k - £30k | 1.02 (0.92, 1.13) | 1.07 (0.94, 1.22) | 1.15 (1.02, 1.3) | 1.43 (1.18, 1.72) | 0.93 (0.83, 1.03) | 0.97 (0.87, 1.08) |
|  | £30k - £60k | 0.91 (0.81, 1.01) | 1.18 (1.03, 1.34) | 1.25 (1.1, 1.43) | 1.89 (1.55, 2.32) | 0.97 (0.87, 1.08) | 0.99 (0.87, 1.11) |
|  | £60k - £90k | 0.8 (0.69, 0.91) | 1.1 (0.93, 1.31) | 1.18 (1, 1.38) | 2.11 (1.58, 2.81) | 0.81 (0.7, 0.93) | 1 (0.87, 1.16) |
|  | £90k+ | 0.88 (0.75, 1.03) | 0.98 (0.81, 1.18) | 0.95 (0.79, 1.15) | 1.71 (1.26, 2.32) | 0.75 (0.64, 0.88) | 0.97 (0.8, 1.17) |
| Living Arrangement (ref. Not alone, no child) | Not alone, with child | 0.93 (0.86, 1.01) | 0.92 (0.84, 1) | 0.94 (0.86, 1.03) | 0.92 (0.79, 1.07) | 0.83 (0.77, 0.9) | 0.87 (0.81, 0.95) |
|  | Alone | 0.83 (0.77, 0.9) | 1.02 (0.93, 1.12) | 1.04 (0.95, 1.14) | 0.88 (0.76, 1.02) | 0.75 (0.69, 0.81) | 0.93 (0.86, 1.02) |
| Household Overcrowding (ref. <1 persons per room) | 1+ person per room | 1.1 (1, 1.21) | 0.84 (0.76, 0.93) | 0.83 (0.75, 0.92) | 0.88 (0.74, 1.04) | 1.2 (1.09, 1.31) | 0.9 (0.82, 0.99) |
|  | Nieghbourhood Crowding | 0.98 (0.9, 1.07) | 0.99 (0.89, 1.1) | 1.03 (0.93, 1.14) | 0.96 (0.79, 1.15) | 0.91 (0.84, 1) | 0.98 (0.9, 1.07) |
| Lockdown 1.0 Mental Health (ref. Same) | Worse | 0.97 (0.88, 1.06) | 0.96 (0.85, 1.07) | 1.01 (0.9, 1.13) | 0.8 (0.67, 0.96) | 1.03 (0.94, 1.14) | 1.01 (0.91, 1.11) |
|  | Better | 0.85 (0.72, 0.99) | 0.87 (0.71, 1.06) | 0.9 (0.73, 1.11) | 0.68 (0.53, 0.89) | 0.99 (0.82, 1.19) | 1 (0.85, 1.19) |
|  | Openness | 1.35 (1.26, 1.44) | 1.2 (1.12, 1.3) | 1.14 (1.06, 1.23) | 1.15 (1.02, 1.3) | 1.28 (1.21, 1.37) | 1.23 (1.15, 1.32) |
|  | Conscientiousness | 1.36 (1.27, 1.45) | 1.15 (1.07, 1.24) | 1.16 (1.08, 1.25) | 0.98 (0.86, 1.1) | 1.45 (1.36, 1.54) | 1.28 (1.2, 1.37) |
|  | Extraversion | 0.91 (0.85, 0.97) | 0.75 (0.69, 0.81) | 0.65 (0.61, 0.7) | 0.85 (0.75, 0.96) | 1.1 (1.03, 1.17) | 0.92 (0.86, 0.98) |
|  | Agreeableness | 1.03 (0.96, 1.1) | 0.92 (0.85, 1) | 0.89 (0.82, 0.96) | 1.26 (1.1, 1.43) | 1.06 (0.99, 1.14) | 1.1 (1.03, 1.19) |
|  | Neuroticism | 0.92 (0.85, 1) | 0.85 (0.77, 0.93) | 0.83 (0.76, 0.91) | 1.19 (1.02, 1.39) | 1.2 (1.11, 1.3) | 0.96 (0.88, 1.05) |
|  | Locus of Control | 0.8 (0.73, 0.87) | 0.88 (0.78, 0.98) | 0.82 (0.74, 0.91) | 0.6 (0.5, 0.72) | 0.78 (0.71, 0.86) | 0.78 (0.7, 0.87) |
|  | Risk Behaviour | 0.49 (0.45, 0.54) | 0.58 (0.52, 0.64) | 0.58 (0.52, 0.64) | 0.46 (0.37, 0.56) | 0.59 (0.54, 0.65) | 0.44 (0.4, 0.49) |
|  | Empathy | 1.48 (1.34, 1.62) | 1.13 (1.01, 1.26) | 1.24 (1.1, 1.39) | 1.65 (1.39, 1.95) | 1.58 (1.44, 1.75) | 1.51 (1.36, 1.67) |
|  | Self-Isolating | 1.62 (1.4, 1.87) | 1.36 (1.12, 1.64) | 1.3 (1.09, 1.56) | 0.81 (0.63, 1.05) | 1.19 (1.03, 1.38) | 1.28 (1.1, 1.5) |
|  | Psychiatric Disorder | 1.06 (0.98, 1.16) | 1.11 (1.01, 1.23) | 1.13 (1.02, 1.25) | 0.87 (0.74, 1.02) | 0.93 (0.85, 1.01) | 1.09 (0.99, 1.19) |
| Long-Term Conditions (ref. 0) | 1 | 1.34 (1.25, 1.44) | 1.21 (1.11, 1.31) | 1.12 (1.04, 1.21) | 0.9 (0.78, 1.03) | 1.2 (1.12, 1.29) | 1.21 (1.12, 1.3) |
|  | 2+ | 1.48 (1.35, 1.63) | 1.29 (1.15, 1.45) | 1.11 (1, 1.24) | 0.79 (0.67, 0.94) | 1.43 (1.3, 1.57) | 1.37 (1.23, 1.51) |
